# An SEIR Model for Assessment of Current COVID-19 Pandemic Situation in the UK

**DOI:** 10.1101/2020.04.12.20062588

**Authors:** Peiliang Sun, Kang Li

## Abstract

The ongoing COVID-19 pandemic spread to the UK in early 2020 with the first few cases being identified in late January. A rapid increase in confirmed cases started in March, and the number of infected people is however unknown, largely due to the rather limited testing scale. A number of reports published so far reveal that the COVID-19 has long incubation period, high fatality ratio and non-specific symptoms, making this novel coronavirus far different from common seasonal influenza. In this note, we present a modified SEIR model which takes into account the time lag effect and probability distribution of model states. Based on the proposed model, it is estimated that the actual total number of infected people by 1 April in the UK might have already exceeded 610,000. Average fatality rates under different assumptions at the beginning of April 2020 are also estimated. Our model also reveals that the *R*_0_ value is between 7.5–9 which is much larger than most of the previously reported values. The proposed model has a potential to be used for assessing future epidemic situations under different intervention strategies.

## 1 Disclaimer

All the data used in the paper are based on publicly available resources, including references to official and professional websites and peer-reviewed journals. The model, data and discussions presented in this paper is for research and education only. The model may not represent the real situation, and it may fail due to inadequate model elements and incorrect initial settings.

## 2 Modelling

The Susceptible-Infectious-Removed (SIR) model, as one of the simplest compartmental models originated in the early 20th century [4], has been widely used to model infectious diseases. Several SIR derivatives which contain more states have been used to predict the COVID-19 pandemic all over the world [1, 3, 6]. In these SIR derivatives, the dynamics of each state is governed by several parameters such as: contact rate, infection rate, and cure rate etc. These averaged parameters are estimated using statistic methods, but their value distributions are rarely taken into account in the SIR modelling, thus the complex dynamics of the COVID-19 may not be captured.

The COVID-19 has shown to have long incubation time, and the time from being infected to recovery/death varies in a large scale. Moreover large proportion of the infected people only show mild symptoms or even have no symptoms, implying that the spread of the virus could be far more broad.

Authors in [9] analysed the probability distribution of onset to death and onset to recover time, and the proportion of all infections that would lead to hospitalisation according the data in China. These statistic data are used in this paper to formulate the time delay of COVID-19 status transition with the probability distribution over time.

Figure 2 illustrates the basic structure of the SEIR model, a SIR derivative, where *S*(*t*) represents the susceptible cases at day *t, E*(*t*) is for the exposed cases at day *t, I*(*t*) stands for the infectious cases at day *t* and *I*^Σ^(*t*) denotes the total infected population at day *t. R*(*t*) and *D*(*t*) represent the cumulative recovered cases and cumulative deaths till day *t* respectively. The total of cases that may need hospital treatment at day *t* is denoted as *H*(*t*), while *Q*(*t*) represents the quarantined cases at day *t*. The time variable is *t* = 1, 2, *…, K* and *K* is the prediction horizon.

**Figure 1:**
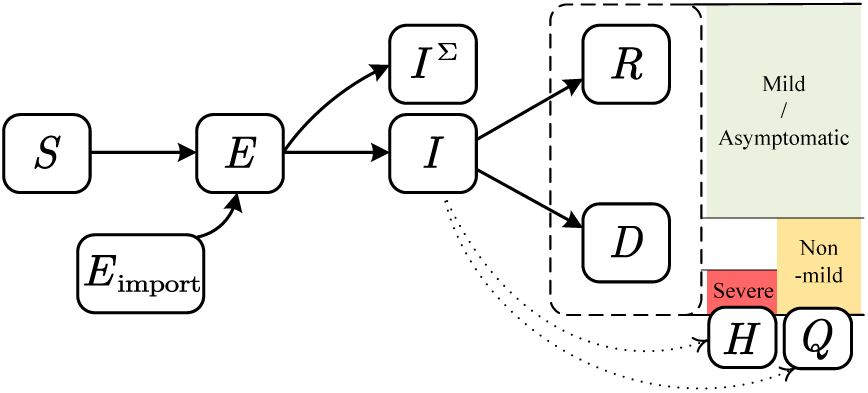
A modified SEIR dynamic model

**Figure 2:**
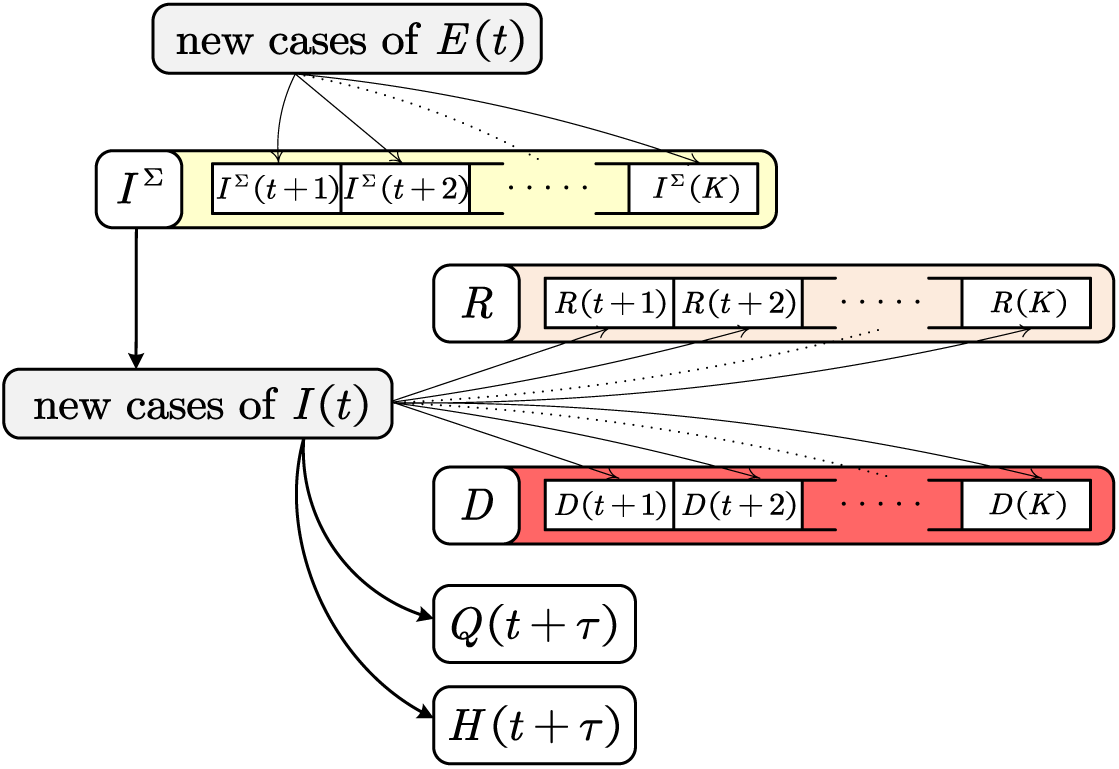
Model state transitions

The model dynamics are formulated as follows: 

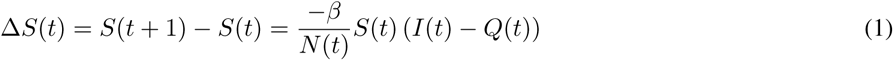

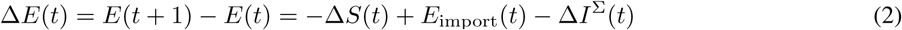

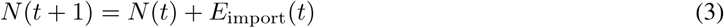

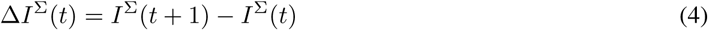

where 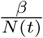 is the contact ratio and *N* (*t*) is the current population in the whole country. In (1), the unquarantined people can contact and spread the virus to the some of the susceptible people Δ*S*(*t*). The imported case *E*_import_(*t*) and Δ*S*(*t*) add up to the increment of *E*(*t*). At each time step a certain proportion of *E*(*t*) will be converted to *I*(*t*) and the increment of total infected people *I*^Σ^(*t*) is subtracted from *E*(*t*) as shown in (2).

However once a person is exposed to the virus and get infected, the virus starts to incubate in his/her body and that person can then become infectious. Therefore a cumulative distribution function of the incubation time in [5] is introduced and is shifted two days^1^ before the symptoms onset to the new cases of *E*(*t*) to *I*^Σ^(*t*) as shown in Figure 2, 

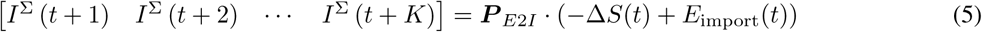

where ***P*** _*E*2*I*_ is a ro w vector of dimension *K* representing the cumulative probability.

Similarly the number of recovered and death cases can also be derived by introducing two separate cumulative probability vectors ***P*** _*I*2*R*_ and ***P*** _*I*2*D*_ as shown in (6) and (7), 

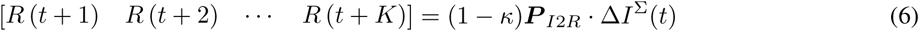

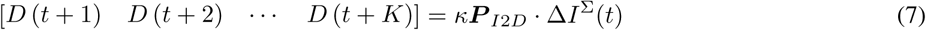

where *κ* is the average infection fatality rate (IFR).

The state *Q*(*t*) is estimated three days^2^ after onset of non-mild symptoms, i.e. after people develop fever/dry cough symptoms they need either be under quarantine in hospital or start self-isolation at home. State *H*(*t*) is used to represent the accumulation of people that need hospital treatment. 

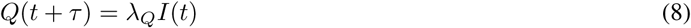

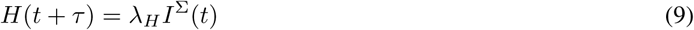

where *τ* is the time period from becoming infectious to having developed non-mild symptoms^3^ that need either admission to hospital or self-isolation at home, *λ*_*Q*_ and *λ*_*H*_ are ratio of non-mild symptoms and ratio of people needing hospital admission.

Finally, the infectious people i.e. state *I* at day *t* can be updated by (10). 

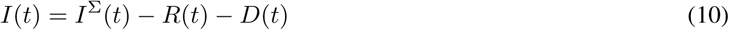

Apparently at day *t, N* (*t*) = *S*(*t*) + *E*(*t*) + *I*(*t*) + *R*(*t*) + *D*(*t*) is automatically satisfied.

## 3 Modelling Assumptions and Parameter Setting

The real epidemiological dynamics is extremely complex and we made several assumptions to set parameters and to simplify the model:

1. The genetic difference in human race is not considered. The cumulative probability functions used in the model are acquired from the analysis of the COVID-19 data from China, and are applied to the UK situation under the assumption that this virus spreading mechanism remains unchanged.
2. About 20% of the infected people will develop symptoms and can be classified as quarantined cases.
3. Infected people can become infectious two days before symptom onset.
4. The virus tests can be conducted to each patient in severe situation on the third day after COVID-19 symptom onset.
5. The officially published death case number is accurate [7].
6. The total confirmed case number is larger than the number of cases that require hospital treatment (severe cases).
7. The earliest imported cases occurred on 15 Jan 2020 [8] and the imported cases kept growing at an exponential rate till international flights were reduced.

**Table 1:** Model Parameters

| Symbol | Description | Value |
| --- | --- | --- |
| $N(1)$ | Initial population | 110 Million |
| $E(1)$ | Initial exposed cases | 5 |
| $\Delta E_{\text{import}}^{\text{max}}$ | Maximum imported cases | 100–10k |
| $\beta_0$ | Infection rate without intervention | 0.15–0.5 |
| $\lambda_Q$ | Ratio of people who have non-mild symptoms | 20% |
| $\lambda_H$ | Ratio of people who need hospital treatment | 1–6% |
| $\kappa$ | Infection fatality ratio | 0.5–3% |
| $\tau$ | Time period from being infectious to having non-mild symptom | 5 days |

Based on published results, official statistics and announcements, the parameters used to build the UK pandemic model are illustrated in Fig. 4 and Fig. 5.

**Figure 4:**
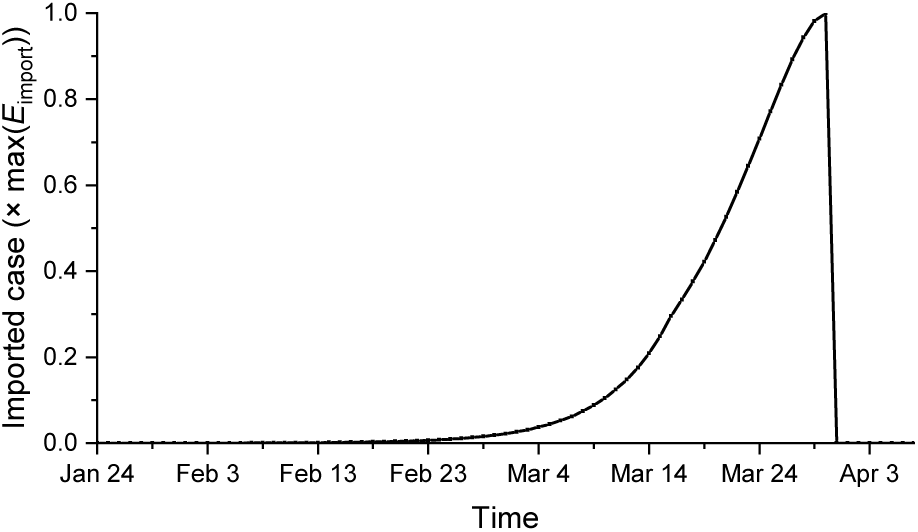
Imported exposed cases

**Figure 5:**
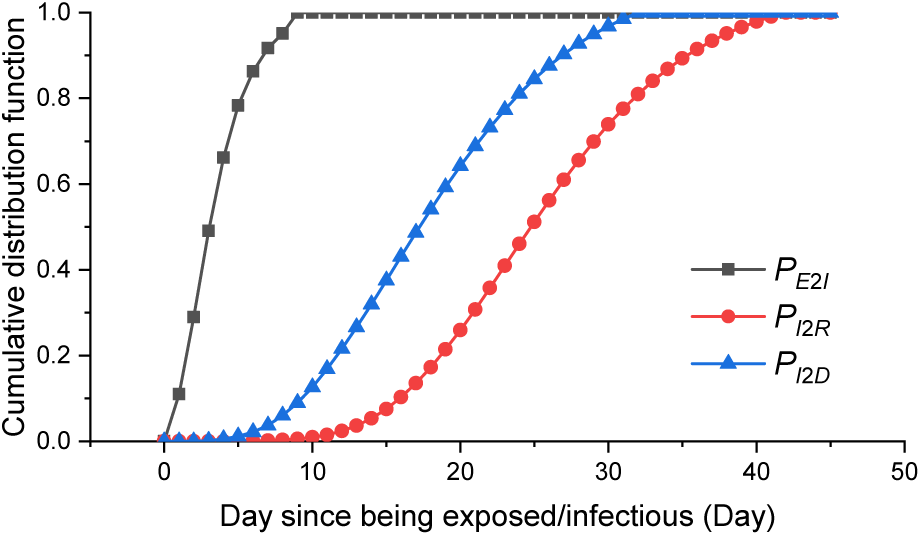
Cumulative distribution functions of ***P*** _*E*2*I*_, ***P*** _*I*2*R*_, ***P*** _*I*2*D*_

By the end of March 2020 international flights are greatly reduced from the normal capacity, we assume the number of imported cases grew exponentially before 16 March. The international flight capacity was decreased linearly afterwards and by the end of March most of the international flights are grounded. The number of imported cases is used as an adjustable parameter and is also used as an initial condition for future projection.

As shown in Figure 3, from 23 March, pubs and schools are closed and people dispersed to their home to work and study. So we simply assume the infection rate^4^ as: 

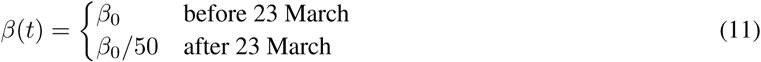

**Figure 3:**
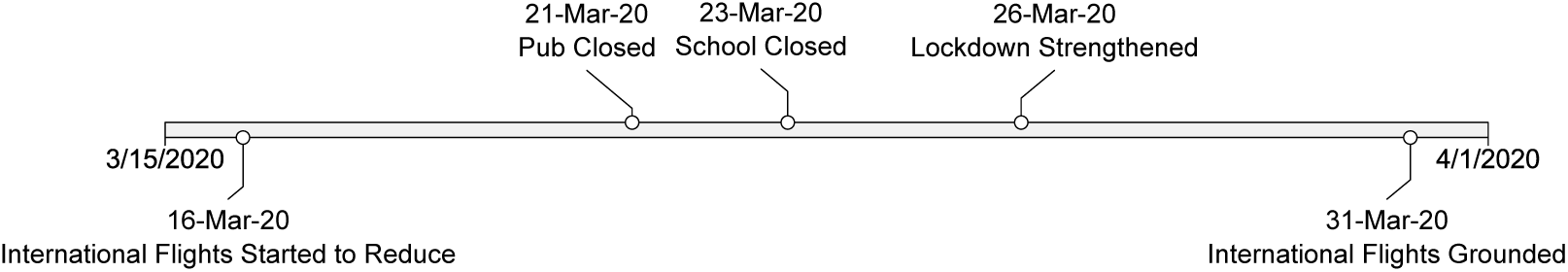
Intervention timeline

The actual infection fatality ratio *κ* and the severe case ratio *λ*_*H*_ can not be accurately calculated, while the reported total case number might be way below the actual infected cases, therefore we have adopted the following procedures to fit the model to the UK death data (1 Feb to 10 Apr):

1. Select a set of *λ*_*H*_ and *κ* satisfying *λ*_*H*_ *> κ*.
2. Search feasible combination of 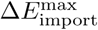 and *β*_0_ such that the following conditions (a)–(c) are met:
  a. The average of daily reported cases must be greater than the average of estimated *H*(*t*) over the whole data fitting period, and the number of reported cases should be greater than the model estimated *H*(*t*) in the last day of the whole period, i.e. 10 April;
  b. The increment of *H*(*t*) on 8 April was lower than the increment of *H*(*t*) on 7 April based on a news report^5^ that the growth in the number of hospitalized coronavirus patients is definitely getting slower;
  c. *D*(*t*) on 10 April should be greater than reported death toll on the same day, i.e. 10 April.
3. The best combination of *β*_0_ and 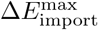 are selected such that the estimated curve of *D*(*t*) fits the daily reported total death number over the data fitting period with the least normalized mean square error.

## 4 Discussions on Nowcasting and Forecasting

### 4.1 Epidemic prevalence estimation on 1st April 2020

By performing the aforementioned modeling procedures, estimates of the number of actual infected people with different infection fatality ratio *κ* and different severe case ratio *λ*_*H*_ are plotted in Figure 6. The figure indicates that although the actual IFR *κ* cannot be calculated accurately, based on our modeling results shown in Figure 6, the total number of infected cases could have been greater than 0.61 million by 1 April 2020. If the IFR *κ* was lower than 2%, the total case number could be even greater than 1 million. Several reports indicate that IFR is below 1% [2], but based on our model, the lower IFR *κ* implies lower severe case ratio *λ*_*H*_ and larger infection number. Hence, the total number of infections would exceed 2 million if IFR *κ* is below 1%. However, according to the UK demographic data, about 5% infected individuals may need hospital treatment, therefore a higher IFR (*κ >* 2%) is more likely to be true in the early stage of this virus outbreak.

**Figure 6:**
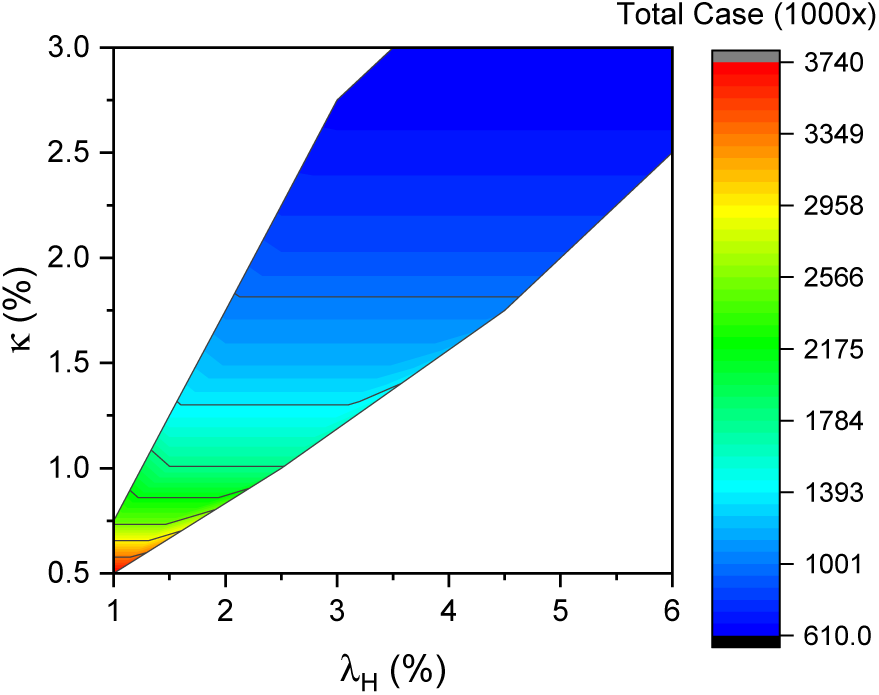
Estimation of the number of actual infected people by 1 April 2020

By curve-fitting the results illustrated in Figure 6, the estimate of the infection rate *β* is around 0.41. The basic reproduction ratio *R*_0_ can be estimated by calculating *S* (*∞*)*− S*(1) while setting *E*(1) = 1, Δ*E*(*t*) = 0. In our estimate *R*_0_*≈* 8.8 which is much larger than the results obtained in other published papers. However if we assume anyone who is infected by the virus will be under quarantine 7 days after s/he becomes infectious, then our model estimate for *R*_0_ *≈* 2.8–3.6 which is comparable to the current popular estimates of *R*_0_ reported in the literature [10]. The reason for our estimated *R*_0_ being much larger than the *R*_0_ estimates reported in the literature might be due to that there is a large proportion of people who only got mild symptoms or even were asymptomatic. They can spread the virus before symptom onset till recovery over a long period of time before the lockdown measure was put in place.

### 4.2 Assessment of future epidemic situation

As presented in the previous section, although the severe case ratio *λ*_*H*_ and the infection fatality ratio *κ* cannot be accurately obtained, our model produces some consistent estimates on the total death toll if community spread is greatly suppressed after 23 March. If the UK government continues the current lockdown strategy to the end of June, the estimated total death number is about 21000 (if the reproduction ratio is suppressed to 0.2). Figure 7 and Figure 8 illustrate the total number of deaths and infections to the end of April if the reproduction ratio is controlled to be less than 0.2 after the lockdown, and assume the severe case ratio *λ*_*H*_ = 4%, infection fatality ratio (IFR) *κ* = 2%, and the maximum imported cases 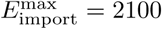. These figures suggest that about 1 million people would be infected, and 81.2% of infected people will be recovered by the end of April in this scenario.

**Figure 7:**
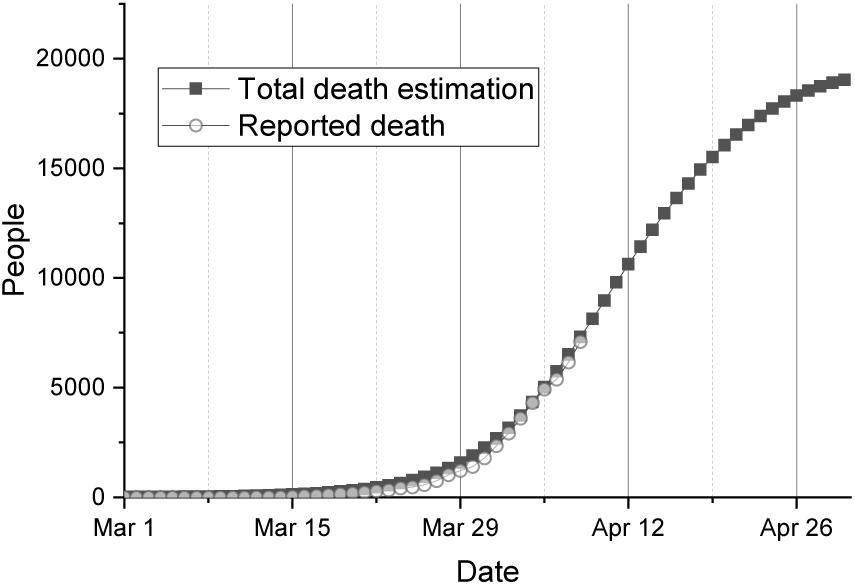
Total death toll projection

**Figure 8:**
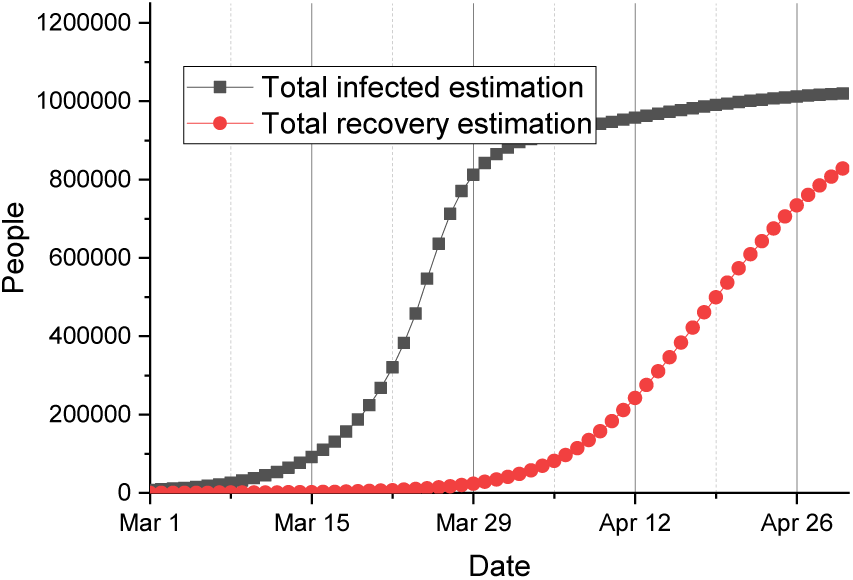
Total infection number projection

However the death toll might be higher if the lockdown rules are not strictly obeyed by people unless improved medical treatment is available in the forthcoming weeks. On the other hand, a long period of lockdown may damage the economy to an unacceptable extent, and the wishes to lift the lockdown measure as early as possible may grow with time.

Figure 9 and Figure 10 show different curves representing possible daily increase in death toll and daily increase in hospital treatment requirements under different reproduction rates. Clearly if the reproduction rate is suppressed below 1, both numbers will keep decreasing. However even the viral transmission is managed to the minimum, the daily death toll will still likely keep at a high level (>500) for two weeks. Based on the current data^6^ in the UK, our model also projects that the inflection point of current pandemic wave in the UK is likely to occur in the following week (between 12-15 April 2020) if the lockdown rules are strictly obeyed by people.

**Figure 9:**
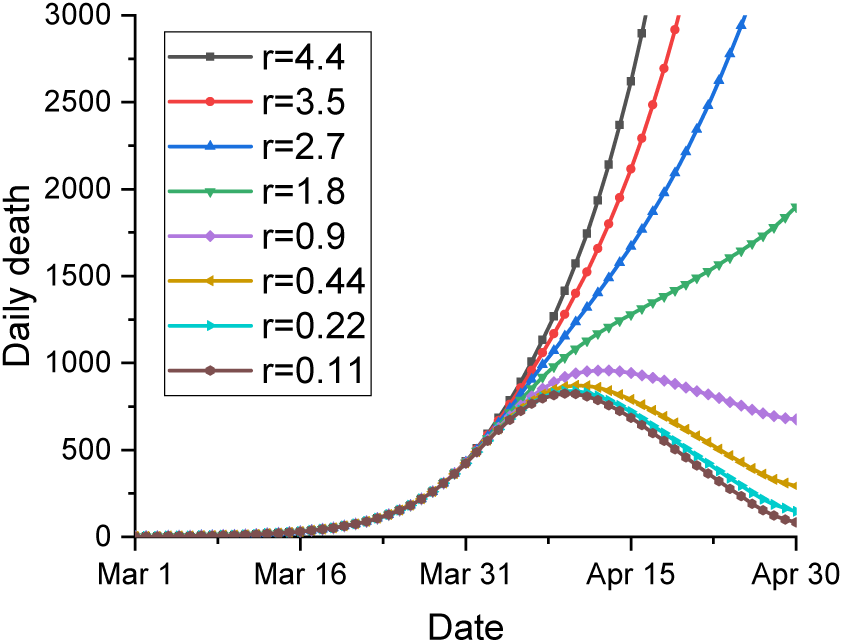
Daily death toll projection

**Figure 10:**
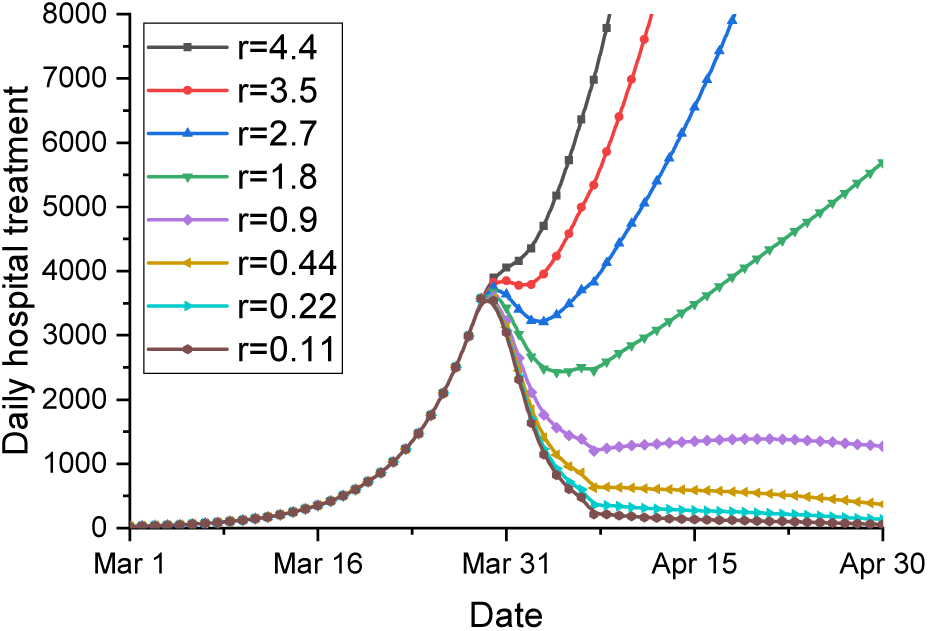
Daily increase in hospital treatment

The model shows that in the ideal situation where everyone stays at home until the end of April, then the daily death number can be suppressed to less than 200 or even less afterwards. Similarly the curve of the number of infected people who need hospital treatment can also be flattened by mid of April.

According to Figure 11^7^, if people start to return to work in May, then the reproduction ratio has to be suppressed below 2 in order that the NHS resources are sufficient to cope with the medical treatment requirements in the next two months (May and June).

**Figure 11:**
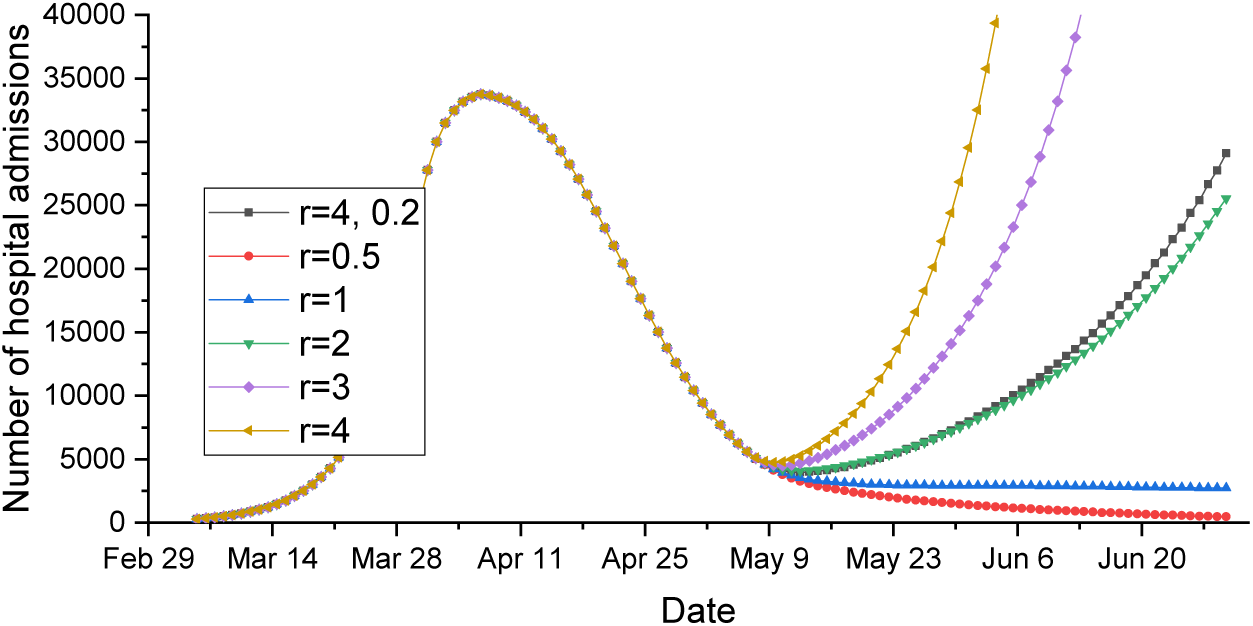
Projection of hospital beds demand for COVID-19

However if the reproduction ratio can not be suppressed well enough while the lockdown measure is lifted in May, one feasible solution to slow down the transmission might be to allow people work every other day. This kind of intervention measures (e.g. only allow people to go outside every other day) has a similar pattern (r=4,0.2)^8^ as the curve with a halved reproduction ratio (r=2).

## 5 Conclusion and Discussion

In this paper, a modified SEIR model has been proposed to allow the state transitions to be governed by the cumulative probability vector rather than by an averaged value, so the COVID-19 data can be utilised to reflect the distribution characteristics over a long period of time. Time variant parameters such as the number of imported cases and infection rate are also introduced and designed to reflect the changes introduced by adopting intervention measures such as reducing the number of international flights and the lockdown measures.

The model is fitted using the officially published data. Because the number of total reported cases may not precisely reflect the reality, the death toll is used instead to assess the pandemic progression in the UK. The results reveal that more than 610,000 people might have been infected by 1 April 2020 and the average death rate in last month is very likely to be greater than 1% and may be even higher than 2%. Also according to the proposed model, the value of *R*_0_ is much greater than the previously published values in the literature, if no intervention measure is implemented. The estimated range of *R*_0_ is between 7.5–9 due to large proportion of patients with mild symptoms. However, if people could keep self-isolated after developing COVID-19 symptoms, then the reproduction ratio is about 2–3, which is comparable to the values published in the literature.

Our model reveals that if people fully comply the ‘stay at home’ order to the end of April, the total death toll is likely to be less than 21,000. To lift the lockdown measure from May, people will need to keep the strict social distance rule and apply other personal protection measures to ensure that the reproduction ratio is suppressed below 2 in order that the NHS has the sufficient capacity to cope with the medical treatment demand from people having severe COVID-19 symptoms. However, if this is not achievable then interventions measures such as people stay at home every other/third day may equally reduce the reproduction ratio in the near future.

The above projections and discussions are based on the assumption that there is no international imported cases after May. However it is likely that the current global pandemic may not reach its end in the next few months, therefore how to limit international imported cases would become a crucial issue to tackle in the future epidemic prevention.

## Data Availability

The data used in the paper is extracted from
The Incubation Period of Coronavirus Disease 2019 (COVID-19) From Publicly Reported Confirmed Cases: Estimation and Application
DOI: 10.7326/M20-0504
Estimates of the severity of coronavirus disease 2019: a model-based analysis
https://doi.org/10.1016/S1473-3099(20)30243-7
and daily reported data from the official reports
https://www.gov.uk/guidance/coronavirus-covid-19-information-for-the-public

## Footnotes

1 According to WHO report people can shed COVID-19 virus 24-48 hours prior to symptom onset. https://www.who.int/docs/default-source/coronaviruse/situation-reports/20200306-sitrep-46-covid-19.pdf?sfvrsn=96b04adf_4

2 In average, patients may need hospital admission on the 3rd day after symptom onset. https://patient.info/news-and-features/coronavirus-how-quickly-do-covid-19-symptoms-develop-and-how-long-do-they-last

3 According to the WHO report, 80% of the patients experienced mild illness https://www.who.int/docs/default-source/coronaviruse/situation-reports/20200301-sitrep-41-covid-19.pdf?sfvrsn=6768306d_2

4 This model is used to fit the death case curve, the variation on *β*(*t*) after lockdown does not affect death number from February to the early April. So for convenience,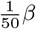is used.

5 https://www.hsj.co.uk/coronavirus/covid-19-hospital-admissions-flattening/7027364.article

6 up to 10 Apr 2020

7 Because the hospitalised number is based on Chinese data. It can be different from the UK scenario where the hospital admission procedures are different. In Figure 11, *λ*_*H*_is set to 4%.

8 When people go out for work the reproduction ratio *r* may become 4. On the next day all people should stay at home and *r* is suppressed to 0.2.

